# Impact of a school-based nutrition education intervention on students’ knowledge, attitudes, and behaviors in Chiang Mai, Thailand: a quasi-experimental study

**DOI:** 10.1101/2025.10.15.25338121

**Authors:** Panrawee Praditsorn, Tina Beuchelt, Ute Nöthlings, Nuttarat Srisangwan, Arisa Keeratichamroen, Kitti Sranacharoenpong, Christian Borgemeister

**Affiliations:** Center for Development Research, University of Bonn, Bonn, Germany; Institute of Nutritional and Food Sciences, Nutritional Epidemiology, University of Bonn, Bonn, Germany; Institute of Nutrition, Mahidol University, Nakhon Pathom, Thailand; ASEAN Institute for Health Development, Mahidol University, Nakhon Pathom, Thailand

**Keywords:** nutrition education, primary school children, knowledge, attitudes, and behaviors, quasi-experimental study, Thailand

## Abstract

The increasing prevalence of childhood obesity in Thailand highlights the need for effective school-based interventions. This study evaluated the impact of a short-term nutrition education intervention on the nutrition knowledge, attitudes, and behaviors (KAB) of students in grades 4-6 in Chiang Mai, Thailand. A quasi-experimental design was implemented in nine public primary schools, comprising four intervention schools (121 students) and five control schools (153 students). The nine-week intervention integrated animated video modules into Health Education, Mathematics, and English lessons. Data on KAB were collected at baseline and at a three-month follow-up after the intervention and analyzed using mixed-effects linear regression models adjusting for clustering at the school level and for student covariates (sex, ethnicity, and daily allowance). Baseline characteristics were comparable between groups, except for age and ethnicity. Knowledge scores increased modestly in both groups, but the difference in change between groups was not significant (adjusted β = 0.21; 95% CI -0.47 to 0.89; p = 0.55). Attitude scores decreased slightly, and behavior scores remained stable, with no significant intervention effects. The short-term, teacher-facilitated intervention modestly improved nutrition knowledge but did not significantly affect overall KAB or dietary patterns. Longer-term, multi-component strategies involving teachers, parents, and supportive school food environments are required to achieve sustained improvements in children’s dietary behaviors.

## Introduction

Childhood obesity has become a public health issue with serious long-term health and socio-economic consequences. Globally, the prevalence of overweight and obese children and adolescents aged 5-19 has risen dramatically from 4% in 1975 to nearly 20% in 2022, a trend observed in both high-income and low- to middle-income countries [1]. Thailand mirrors this trend, with national surveys revealing growing rates. According to the Thai National Health Examination Survey (2019-2020), the prevalence of overweight and obesity among children aged 6-14 years increased 2.7 times from 1996 to 2020, while the rates of underweight, stunting, and thinness declined [2].

Socioeconomic factors influence obesity rates, with higher prevalence observed among Thai children from higher socioeconomic backgrounds [3]. Urbanization and lifestyle changes, such as increased sedentary behavior and consumption of energy-dense, nutrient-poor foods, have exacerbated these trends [4]. Research conducted in Thailand has shown that urban life is associated with an increased risk of obesity [5]. Chiang Mai, the largest city in northern Thailand, reflects this trend due to rapid urbanization and socio-economic development, which has contributed to rising childhood obesity rates [5, 6].

To address this issue, school nutrition education has gained recognition as an effective strategy to improve dietary behaviors and prevent obesity in children. A systematic review found that school-based interventions targeting dietary and physical activity behaviors successfully promoted healthier eating and body weight management. The involvement of multiple stakeholders and the integration of educational activities into school curricula were critical to these interventions’ success [7]. Another review highlighted that many school-based programs targeting dietary behaviors and physical activity positively impacted childhood obesity prevention, with teachers playing a key role in delivering the curriculum [8]. In the U.S., school-based nutrition education programs were shown to positively influence students’ dietary choices, encouraging healthier eating behaviors [9]. Similarly, schools in China employing health-promoting frameworks demonstrated significant improvements in both nutrition knowledge and eating behaviors among students who received basic health education [10].

Thailand has undergone a nutritional transition, shifting from primarily addressing undernutrition to now confronting a dual burden of undernutrition and rising obesity. National programs, emphasizing nutrition education and public health campaigns, have been instrumental in this endeavor. School-based interventions have demonstrated effectiveness in promoting healthy eating habits among Thai students, particularly increasing fruit and vegetable consumption [11]. However, inconsistent implementation and limited evaluation have hindered a comprehensive understanding of their long-term effectiveness. The continuing rise in childhood obesity underscores the need for further research to assess the impact of these programs and identify opportunities for improvement [12].

This study adapted the nutrition education modules from a previous study in 2018 in Thailand [13]. The earlier study’s limitations included challenges in developing age-appropriate nutrition education tools, with content that was difficult for students in grades 1-3 to understand. In addition, inconsistent timelines across subjects, external factors like reduced learning time due to school activities, and unsuitable classroom setups hindered effective implementation. This highlights the need for a more flexible and integrated approach to nutrition education.

Therefore, this study aimed to evaluate the impact of a school-based nutrition education intervention on the nutrition knowledge, attitudes, and behaviors of students in grades 4-6 attending public primary schools in Chiang Mai, Thailand. The primary objective was to assess the short-term impact of the intervention on nutrition knowledge, with secondary objectives focusing on changes in nutrition-related attitudes and behaviors. It was hypothesized that the intervention would primarily improve students’ nutrition knowledge, with potential positive effects on attitudes and behaviors.

## Methods

### Study Timeline

The research team coordinated with the Chiang Mai Primary Educational Service Area Office 4 and contacted the selected schools between July and September 2022 to present the intervention and obtain administrative and teacher agreement. Baseline data collection was conducted from June to July 2023. Teachers were trained in June 2023 to integrate the nutrition education modules before implementation. The nine-week intervention was delivered from August to September 2023, and post-intervention data were collected three months later, between November 2023 and January 2024.

### Study design and setting

This study employed a cluster quasi-experimental design, using schools as clusters and students as the units of observation. Two districts, Saraphi and Hang Dong, were initially selected because of their similar semi-urban context, comparable population sizes [14], and inclusion under the same administrative authority, the Chiang Mai Primary Educational Service Area Office 4. The districts also exhibit comparable demographic and socioeconomic profiles [15]. Both districts had similar educational infrastructure, and our school sampling strategy ensured comparable representation of small, medium, and large public primary schools. Saraphi was designated as the intervention district and Hang Dong as the control district.

Although schools were randomly selected within each district, the districts themselves were purposively assigned to the intervention or control arm. Consequently, the design is quasi-experimental rather than a true cluster-randomized trial, and district-level confounding cannot be excluded given the small number of districts (n = 2). To minimize bias arising from the hierarchical data structure, all outcomes were analyzed with mixed-effects models with random intercepts for schools. This random-effects approach accounts for correlation among students within the same school and, even with only nine school clusters, provides unbiased estimates of fixed (treatment) effects, although variance components cannot be estimated precisely [16].

The study population was students enrolled in grades 4-6. Prior to data collection, approval was obtained from the Chiang Mai Primary Educational Service Area Office 4, and written informed consent was collected from parents or caregivers of participating students.

### Sampling Method

A multi-stage sampling design incorporating cluster sampling and disproportionate stratified random sampling was used. Initially, Chiang Mai province was purposively selected for cluster sampling due to its rapid urbanization and ongoing nutritional transition, which have resulted in an increasing prevalence of childhood obesity and related dietary issues [5]. After selecting and assigning districts to intervention and control groups, five public primary schools were randomly chosen from each district using disproportionate stratified random sampling based on school sizes.

According to the Ministry of Education, Thailand (2019) [17], schools were classified based on total student enrollment from kindergarten through sixth grade into three categories: small (<121 students), medium (121–280 students), and large (>280 students). Enrollment data were obtained from an officer at the Chiang Mai Primary Educational Service Area Office 4 in May 2023. In Saraphi, the sample included two small, two medium, and one large school, while in Hang Dong, one small, three medium, and one large school were selected. This sampling strategy ensured varied representation across school sizes and supported fair comparisons between intervention and control groups.

### Participants

The study population comprised students in grades 4-6 in public primary schools, as this age group is vulnerable to environmental influences and has relatively easy access to less healthy food in the school neighborhood food environment [18]. The target sample size of 300 students (150 per group) was calculated using G*Power 3.1. Parameters were set to detect a small-to-moderate effect size (Cohen’s d = 0.20) for differences between two independent groups, with a significance level of 0.05 and a statistical power of 0.90. The total sample size included an allowance for a 10% dropout rate. A comprehensive recruitment strategy was implemented to enroll eligible students. A sampling frame was developed using school records, which included each student’s birthdate, weight, and height. Participants were then selected through stratified sampling based on sex (male/female) and nutritional status. Nutritional status was classified according to the Thai Growth Reference (16) into six categories: thinness (<−2SD), mild thinness (−1.5SD to ≥−2SD), normal (−1.5SD to ≤+1.5SD), mild overweight (>+1.5SD to ≤+2SD), overweight (>+2SD to ≤+3SD), and obese (>+3SD). This stratification ensured a balanced representation of students across nutritional and demographic categories. Students with severe cognitive or communication impairments that hindered their participation in the study interviews were excluded from data collection, but continued participating in the nutrition education classes. Exclusion criteria included autism spectrum disorder with severe language or social deficits, intellectual disabilities with marked cognitive impairments, and severe emotional or behavioral disorders that interfere with participation.

### Implementation of nutrition education modules

The intervention was designed to promote healthy eating habits as a strategy to prevent childhood obesity, rather than focusing solely on obesity itself. The nutrition education modules were adapted from those developed by Mongkolsucharitkul et al. (2018) through a collaborative process involving qualitative research, literature review, and input from the Thai research team (13), in alignment with Thailand’s Basic Education Core Curriculum (17). The modules were initially intended for students in grades 1–3, but were considered too advanced for that age group. Therefore, they were adapted for students in grades 4–6.

Each module consisted of 3- to 4-minute animated videos integrated into Health Education, Mathematics, and English lessons to increase engagement and relevance. A blended learning approach was used: video materials were distributed via a Facebook-based e-learning platform, and physical copies with supplementary materials (worksheets, knowledge sheets, and a teaching manual) were also provided to schools.

In June 2023, prior to implementation, a researcher visited each school to provide in-person guidance to the teacher responsible for delivering the nutrition education modules. The one-hour training session introduced the e-learning platform and demonstrated how to integrate worksheets into lessons. Although the intervention was linked to three subjects, Health Education teachers for grades 4-6 were designated as the main facilitators. These teachers, already familiar with the students and curriculum, supported the integration of the program into routine instruction.

The intervention was implemented as a nine-week program comprising three modules, each with three lessons. Teachers were the primary facilitators, delivering the content and providing worksheets for students to complete after each lesson. At the end of each module, a researcher visited each school to summarize the content with the teacher and students. **Table 1** provides a detailed overview of the nutrition education modules.

**Table 1.**
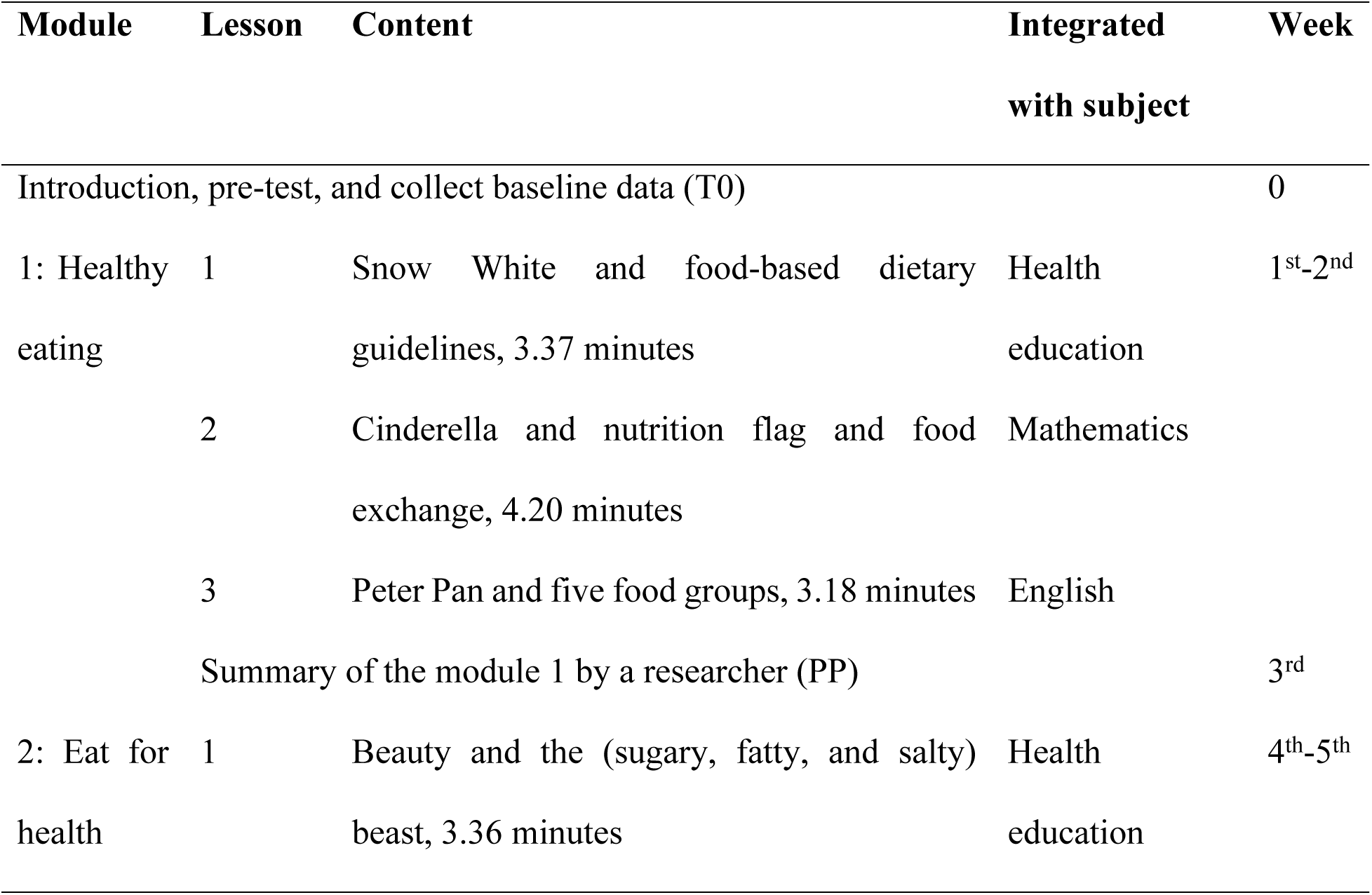
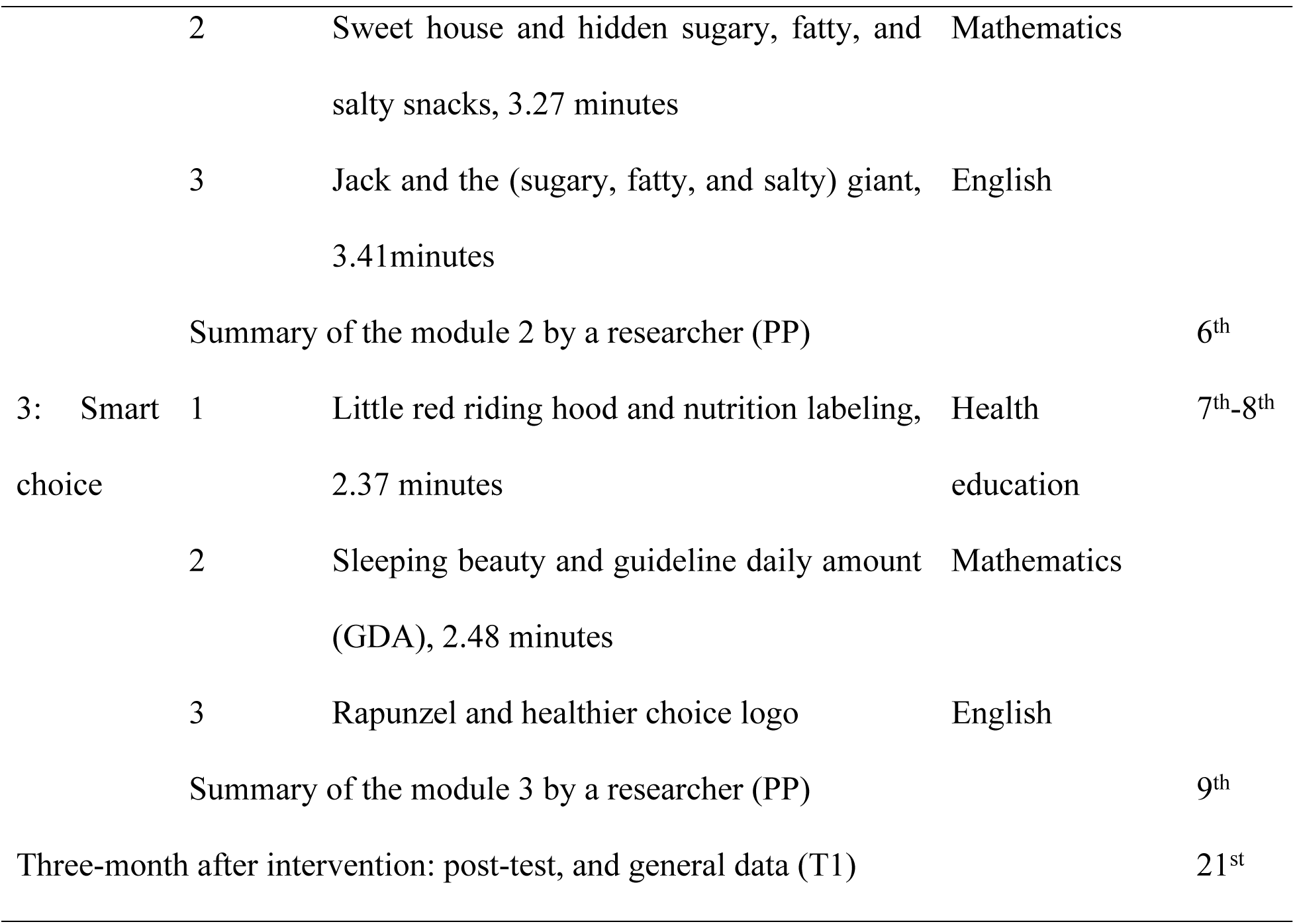
The nine-week nutrition education modules for 4th-6th graders (adapted from (13)).

### Measures

To evaluate the impact of the nutrition education modules, questionnaires were developed to gather data on students’ general information, dietary intake, knowledge, attitudes, and behaviors (KAB), as well as information about the primary household food preparer in both control and intervention groups. Researchers conducted in-person interviews with the students at their schools, except for the household primary food preparer questionnaire, which was completed at home. Each student interview lasted between 30 and 45 minutes.

#### Students’ demographic information

Baseline information included the students’ birthdate, sex, weight, height, and daily allowance. Although school records were initially used to classify students by nutritional status for stratified sampling, these data were not used in the final analysis. Students’ height and weight were re-measured at baseline by the same trained researcher using standardized procedures. Weight was measured using a Tanita HD 380 digital scale, and height was measured using a measuring tape mounted on the classroom wall. This setup remained available for subsequent use by teachers but was initially calibrated and applied by the researcher during baseline data collection. Nutritional status was then determined based on these standardized measurements using the Thai Growth Reference (2019) (16).

#### Household primary food preparer information

At baseline, information regarding the primary parent or caregiver responsible for preparing food at home was collected through a self-reported questionnaire. This included sex, age, weight, height, education, occupation, income, and food preparation practices. Students brought the questionnaire home and returned it the following day. The nutritional status of parents or caregivers was classified based on the World Health Organization (WHO) standards for Asian adults (18).

#### Dietary intake assessment

Dietary intake was assessed at baseline and three months post-intervention using two methods. First, a 24-hour dietary recall (24HDR) captured all foods and beverages consumed the previous day, including portion sizes, which were estimated using food photos, standard utensils (measuring cups, teaspoons, tablespoons, rice spoons), and items available from school shops. Second, a semi-quantitative food frequency questionnaire (semi-FFQ) assessed the frequency of food consumption over the past seven days. The FFQ included the top five items from each food group based on national consumption data for individuals aged ≥5 years (19). Portion sizes were determined by weighing foods purchased locally; for unavailable items, data were obtained from the Thai Food Consumption Table developed by the Institute of Nutrition, Mahidol University (20). The semi-FFQ included 63 items across six food groups and was validated by three experts, achieving a scale-level content validity index of 0.9.

#### KAB test

A KAB questionnaire, initially developed in a previous study (13), was used in this study to align with the content of the nutrition education modules. The questions consisted of 30 items in total: 10 multiple-choice questions assessing nutritional knowledge, and 10 yes/no questions each assessing attitudes toward healthy eating and dietary behaviors. The KAB test was administered before the intervention and three months after its completion. The primary outcome was a change in nutritional knowledge, while secondary outcomes were changes in attitudes and behaviors.

### Ethical approval

The Center for Development Research (ZEF) at the University of Bonn, Germany, approved the study protocol. The approval documents were translated into Thai and submitted to the Chiang Mai Primary Educational Service Area Office 4, which authorized school participation. Information sheets and consent forms for parents and students were provided to teachers at each school, who were instructed to distribute the forms to parents and return the signed forms to the researchers.

### Data analysis

Data were analyzed using STATA version 16 (StataCorp, College Station, Texas, USA). Statistical significance was set at p < 0.05. Descriptive statistics were presented as n (%) for categorical variables, and as mean (SD) or median (P25, P75) for continuous variables. Normality was assessed by the Kolmogorov-Smirnov test and visual inspection of histograms.

Intervention effects were evaluated using mixed-effects linear regression models. Each model included fixed effects for group (intervention vs. control), time (pre- vs. post-intervention), and their interaction (group × time), which tested whether changes over time differed between groups. Random intercepts for school were included to account for clustering of students within schools.

For each outcome (knowledge score, attitude score, and behavior score), both unadjusted and adjusted models were estimated. Adjusted models controlled for a priori students’ covariates, including sex, ethnicity, and daily allowance. Additional sensitivity analyses, adjusting for parental factors and dietary intake, were conducted and are presented in the supplementary tables. The main effect of interest was the group × time interaction, reported as a regression coefficient, 95% confidence interval, and p-value. District-level effects were not modeled due to the small number of districts (n = 2). Missing data were handled by maximum likelihood estimation under the missing-at-random assumption; no imputation was performed. Multicollinearity among covariates was assessed using variance inflation factors (all < 5), indicating no problematic correlations.

## Results

Ten schools were initially selected, with five assigned to the control group (n = 153 students) and five to the intervention group (n = 151 students), totaling 304 students in grades 4-6. However, one intervention school withdrew, excluding 30 students, resulting in five schools in the control group (n = 153 students) and four in the intervention group (n = 121 students). During the nine-week intervention, some students dropped out due to absenteeism or transfers. Post-intervention assessments, conducted three months later, included 145 students in the control group and 120 in the intervention group.

Baseline characteristics of students are presented in Table 2. The two groups were similar in most demographic and anthropometric characteristics. However, students in the control group were slightly older than those in the intervention group (p = 0.003). The distribution of ethnicity differed significantly between groups, with a higher proportion of Thai students in the intervention group and more Tai Yai students, an ethnic minority group originating from Myanmar, in the control group (p = 0.021).

**Table 2.**
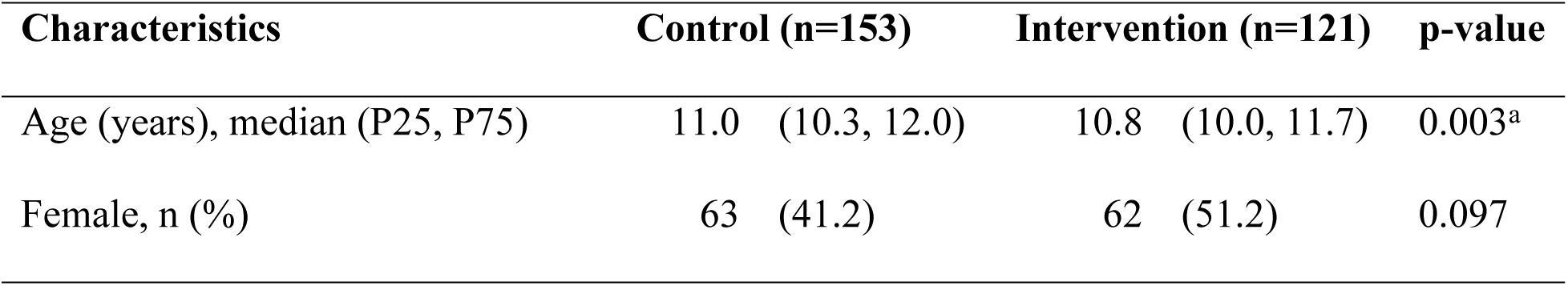
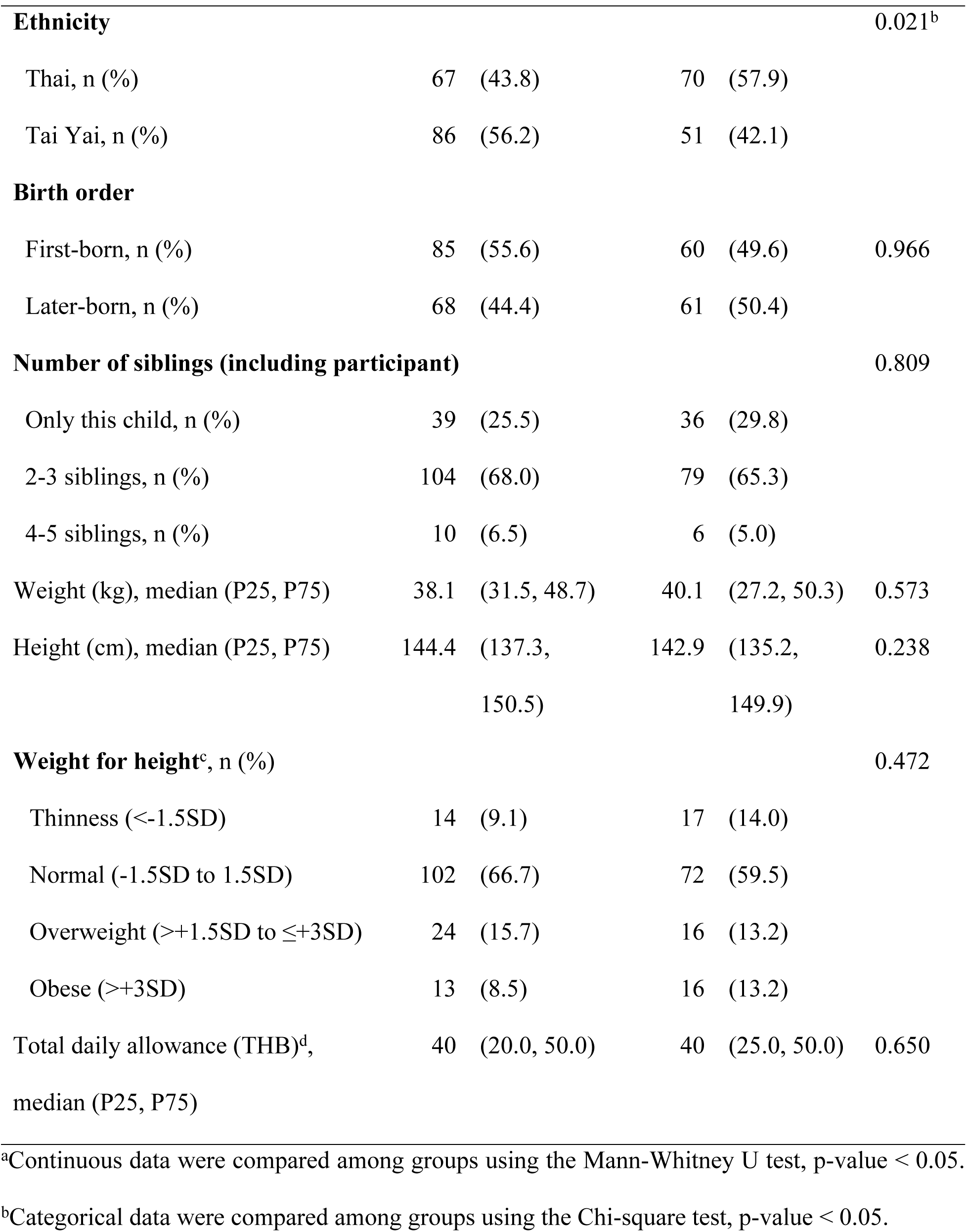
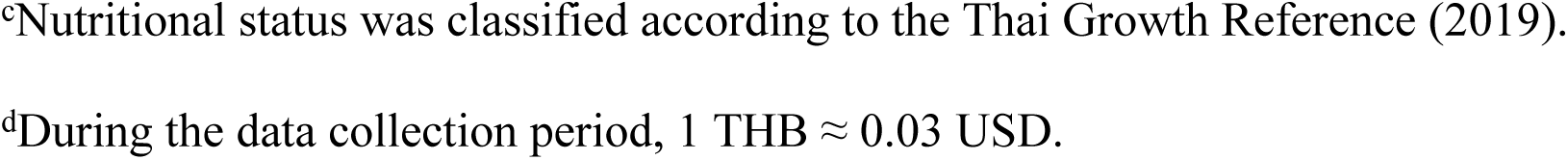
Baseline information of students.

| Characteristics | Control (n=153) |  | Intervention (n=121) |  | p-value |
| --- | --- | --- | --- | --- | --- |
| Age (years), median (P25, P75) | 11.0 | (10.3, 12.0) | 10.8 | (10.0, 11.7) | 0.003<sup>a</sup> |
| Female, n (%) | 63 | (41.2) | 62 | (51.2) | 0.097 |

| Characteristics | Control (n=153) | Intervention (n=121) | p-value |
| --- | --- | --- | --- |
| **Ethnicity** |  |  | 0.021<sup>b</sup> |
| Thai, n (%) | 67 (43.8) | 70 (57.9) |  |
| Tai Yai, n (%) | 86 (56.2) | 51 (42.1) |  |
| **Birth order** |  |  |  |
| First-born, n (%) | 85 (55.6) | 60 (49.6) | 0.966 |
| Later-born, n (%) | 68 (44.4) | 61 (50.4) |  |
| **Number of siblings (including participant)** |  |  | 0.809 |
| Only this child, n (%) | 39 (25.5) | 36 (29.8) |  |
| 2-3 siblings, n (%) | 104 (68.0) | 79 (65.3) |  |
| 4-5 siblings, n (%) | 10 (6.5) | 6 (5.0) |  |
| Weight (kg), median (P25, P75) | 38.1 (31.5, 48.7) | 40.1 (27.2, 50.3) | 0.573 |
| Height (cm), median (P25, P75) | 144.4 (137.3, 150.5) | 142.9 (135.2, 149.9) | 0.238 |
| **Weight for height<sup>c</sup>, n (%)** |  |  | 0.472 |
| Thinness (<-1.5SD) | 14 (9.1) | 17 (14.0) |  |
| Normal (-1.5SD to 1.5SD) | 102 (66.7) | 72 (59.5) |  |
| Overweight (>+1.5SD to ≤+3SD) | 24 (15.7) | 16 (13.2) |  |
| Obese (>+3SD) | 13 (8.5) | 16 (13.2) |  |
| Total daily allowance (THB)<sup>d</sup>, median (P25, P75) | 40 (20.0, 50.0) | 40 (25.0, 50.0) | 0.650 |

<sup>a</sup>Continuous data were compared among groups using the Mann-Whitney U test, p-value < 0.05.
<sup>b</sup>Categorical data were compared among groups using the Chi-square test, p-value < 0.05.
<sup>c</sup>Nutritional status was classified according to the Thai Growth Reference (2019).
<sup>d</sup>During the data collection period, 1 THB ≈ 0.03 USD.

At baseline and post-intervention, mean nutrition knowledge, attitude, and behavior (KAB) scores were similar between the intervention and control groups across the overall sample and when stratified by sex and ethnicity (Table 3). In both groups, total nutrition knowledge scores remained below 50% of the maximum possible, with modest increases after the intervention. Attitude and behavior scores also showed minimal between-group differences at both time points. The only statistically significant difference was observed at baseline, where girls in the intervention group had slightly higher behavior scores than those in the control group; no other subgroup differences were apparent.

**Table 3.** Mean (SD) nutrition KAB scores and between-group comparisons.

| Variables | Baseline |  | p-value | Post-intervention |  | p-value |
| --- | --- | --- | --- | --- | --- | --- |
|  | Control | Intervention |  | Control | Intervention |  |
|  | (n = 153) | (n = 121) |  | (n = 145) | (n = 120) |  |
| Knowledge score (total = 15) |  |  |  |  |  |  |
| Overall | 6.6 (1.9) | 6.5 (2.1) | 0.755 | 7.0 (2.1) | 7.1 (2.1) | 0.591 |
| Gender |  |  |  |  |  |  |
| Male | 6.5 (1.9) | 6.5 (2.1) | 0.954 | 6.7 (2.1) | 6.9 (2.3) | 0.652 |
| Female | 6.7 (1.9) | 6.5 (2.1) | 0.551 | 7.3 (2.2) | 7.3 (1.9) | 0.965 |
| Ethnicity |  |  |  |  |  |  |
| Thai | 6.8 (1.8) | 6.3 (2.1) | 0.098 | 6.8 (1.8) | 6.7 (1.9) | 0.722 |
| Tai Yai | 6.4 (2.0) | 6.8 (2.1) | 0.238 | 7.1 (2.3) | 7.7 (2.2) | 0.160 |
|  | Control | Intervention |  | Control | Intervention |  |
|  | (n = 153) | (n = 121) |  | (n = 145) | (n = 120) |  |
| Attitude score (total = 10) |  |  |  |  |  |  |
| Overall | 5.8 (1.9) | 6.0 (1.8) | 0.388 | 5.5 (2.1) | 5.4 (2.1) | 0.567 |
| Gender |  |  |  |  |  |  |
| Male | 6.0 (2.1) | 5.9 (1.8) | 0.770 | 5.6 (2.2) | 5.2 (2.3) | 0.293 |
| Female | 5.7 (1.7) | 6.2 (1.8) | 0.098 | 5.4 (1.9) | 5.5 (1.9) | 0.726 |
| Ethnicity |  |  |  |  |  |  |
| Thai | 5.8 (1.9) | 6.1 (1.7) | 0.331 | 5.6 (2.1) | 5.3 (2.2) | 0.333 |
| Tai Yai | 5.9 (2.0) | 6.0 (2.0) | 0.766 | 5.4 (2.0) | 5.5 (1.9) | 0.801 |
| Behavior score (total = 10) |  |  |  |  |  |  |
| Overall | 5.8 (1.8) | 6.0 (1.8) | 0.372 | 5.6 (1.8) | 5.9 (1.9) | 0.179 |
| Gender |  |  |  |  |  |  |
| Male | 6.0 (1.9) | 5.8 (1.7) | 0.492 | 5.7 (1.9) | 5.9 (1.9) | 0.452 |
| Female | 5.6 (1.7) | 6.3 (1.9) | 0.048 | 5.4 (1.8) | 5.8 (1.8) | 0.200 |
| Ethnicity |  |  |  |  |  |  |
| Thai | 5.7 (1.7) | 6.0 (1.8) | 0.363 | 5.8 (1.8) | 6.0 (1.9) | 0.435 |
| Tai Yai | 5.9 (1.9) | 6.1 (1.9) | 0.644 | 5.4 (1.9) | 5.6 (1.8) | 0.436 |

P-values are from independent-samples t-tests comparing control and intervention groups at each time point within the subgroup. n = number of students.

Mixed-effects regression analyses (Table 4) showed no significant group × time interactions for KAB scores in either the unadjusted (Model 1) or adjusted models (Model 2). Knowledge scores increased modestly from baseline to post-intervention in both groups, but the difference in change between groups was not significant (adjusted β = 0.21; 95% CI –0.47 to 0.89; p = 0.55). Attitude scores declined slightly (β = –0.39; 95% CI –1.05 to 0.27; p = 0.25), and behavior scores remained essentially unchanged (β = 0.01; 95% CI –0.60 to 0.62; p = 0.97). Overall, these findings indicate that the nine-week nutrition education intervention did not produce measurable improvements in students’ KAB after adjustment for clustering at the school level and individual covariates.

**Table 4.**
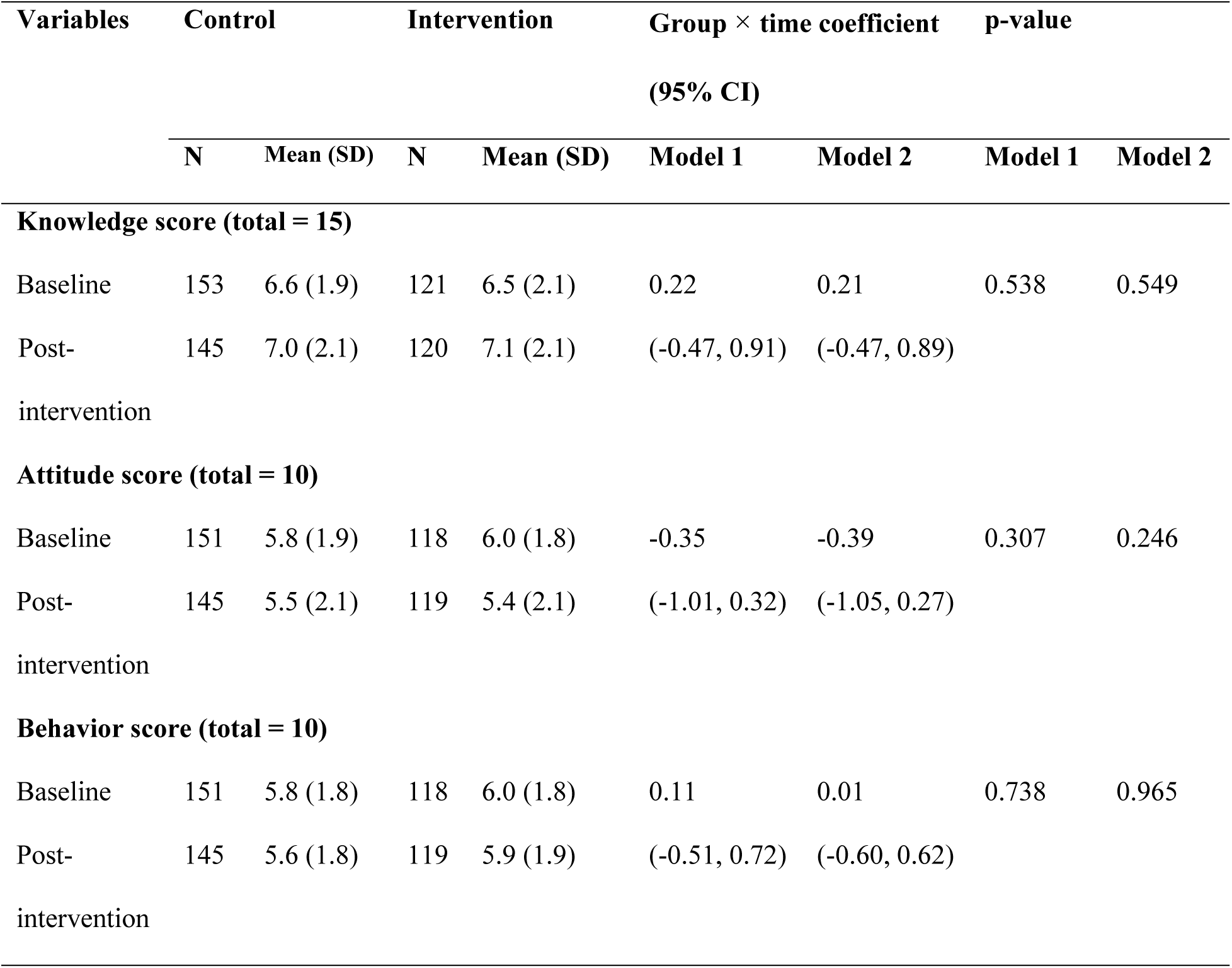
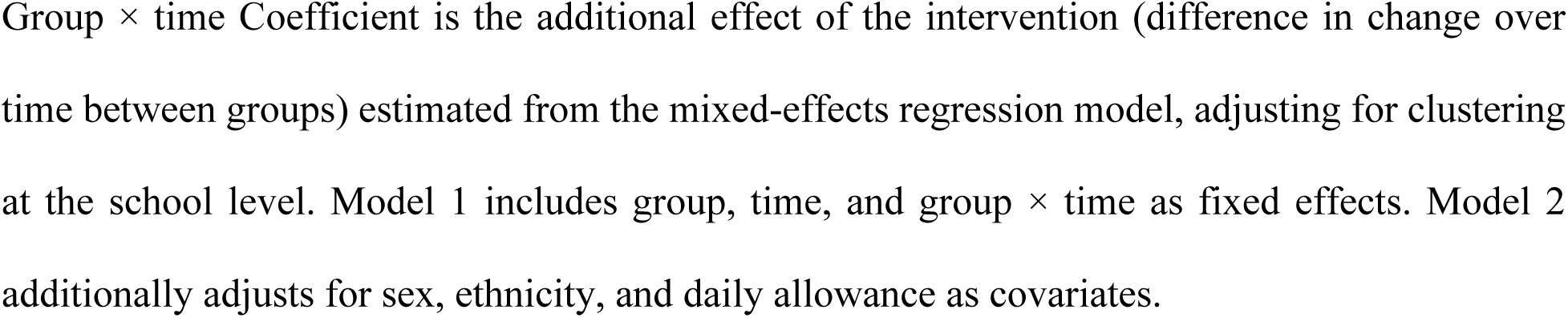
Effectiveness of nutrition education intervention on KAB scores.

| Variables | Control |  | Intervention |  | Group × time coefficient<br>(95% CI) |  | p-value |  |
| --- | --- | --- | --- | --- | --- | --- | --- | --- |
|  | N | Mean (SD) | N | Mean (SD) | Model 1 | Model 2 | Model 1 | Model 2 |
| Knowledge score (total = 15) |  |  |  |  |  |  |  |  |
| Baseline | 153 | 6.6 (1.9) | 121 | 6.5 (2.1) | 0.22 | 0.21 | 0.538 | 0.549 |
| Post-intervention | 145 | 7.0 (2.1) | 120 | 7.1 (2.1) | (-0.47, 0.91) | (-0.47, 0.89) |  |  |
| Attitude score (total = 10) |  |  |  |  |  |  |  |  |
| Baseline | 151 | 5.8 (1.9) | 118 | 6.0 (1.8) | -0.35 | -0.39 | 0.307 | 0.246 |
| Post-intervention | 145 | 5.5 (2.1) | 119 | 5.4 (2.1) | (-1.01, 0.32) | (-1.05, 0.27) |  |  |
| Behavior score (total = 10) |  |  |  |  |  |  |  |  |
| Baseline | 151 | 5.8 (1.8) | 118 | 6.0 (1.8) | 0.11 | 0.01 | 0.738 | 0.965 |
| Post-intervention | 145 | 5.6 (1.8) | 119 | 5.9 (1.9) | (-0.51, 0.72) | (-0.60, 0.62) |  |  |

Group × time Coefficient is the additional effect of the intervention (difference in change over time between groups) estimated from the mixed-effects regression model, adjusting for clustering at the school level. Model 1 includes group, time, and group × time as fixed effects. Model 2 additionally adjusts for sex, ethnicity, and daily allowance as covariates.

At baseline, most household food preparers were mothers and in their late thirties (S1 Table). The majority had normal BMI, primary-level education, and worked in the informal sector. Home-cooked meals were typical. S2 Table shows that students’ energy and nutrient intakes were below Thai Dietary Reference Intake (DRI) recommendations at both time points. Fat, sodium, and sugar intakes exceeded recommended levels, while micronutrient intakes, particularly calcium, iron, zinc, and vitamins A and C, were inadequate. Food frequency data (S3 Table) indicated broadly consistent dietary patterns across groups and time points. Rice, noodles, and fried foods were consumed most frequently, while fruit and vegetable intake remained low. Sugary drinks and snack foods were commonly reported in both groups.

Including parental factors (relationship to the child, education, occupation, income, age, sex, BMI, and food preparation behavior) in mixed-effects models (S4 Table) did not change the findings. No significant group × time interactions were observed for knowledge, attitude, or behavior scores. Additional adjustment for energy and nutrient intakes (S5 Table) and for food-group variables from the semi-quantitative FFQ (S6 Table) also yielded similar results.

Across all supplementary analyses, results were consistent with the primary models, showing no significant intervention effects on students’ nutrition knowledge, attitudes, or behaviors.

## Discussion

This study evaluated the effect of a nine-week school-based nutrition education intervention on the nutrition knowledge, attitudes, and behaviors (KAB) of primary school students in Chiang Mai, Thailand. The findings showed no significant differences between the intervention and control groups in changes to KAB scores. Although knowledge scores increased modestly in both groups, attitude and behavior scores remained unchanged. These results suggest that a short-term intervention focusing primarily on classroom-based education is insufficient to produce measurable improvements in students’ dietary behaviors.

Our findings are consistent with previous studies indicating that gains in nutrition knowledge do not necessarily translate into behavioral change among children. Short-term interventions often improve awareness but fail to influence daily food choices without reinforcement from the home or school environment. Similar findings have been reported in studies from the United States, the United Kingdom, and Southeast Asia, where brief nutrition education programs improved knowledge but not long-term eating habits [19-21]. In contrast, multi-component or longer-term interventions that combine education with environmental and parental involvement have shown stronger effects on children’s eating behavior and body weight outcomes [22].

Some baseline differences were observed between the intervention and control groups, particularly in students’ age and ethnicity distributions. The average age difference was small (11.0 vs. 10.8 years) and unlikely to have influenced outcomes, as all participants were in grades 4–6. Given this narrow age range, age was not included as a covariate in the mixed-effects models. Ethnicity and daily allowance were included as potential confounders, although residual confounding cannot be completely ruled out. The withdrawal of one intervention school reduced the sample size, potentially limiting statistical power, while the small number of school clusters constrained the precision of random-effects estimates. Nevertheless, both districts shared similar semi-urban contexts, and students’ dietary patterns and household characteristics were comparable at baseline, minimizing the likelihood of systematic bias.

The descriptive dietary data provide valuable context for understanding the challenges facing Thai schoolchildren. Across both groups, fruit and vegetable intake was low, while sugary and fried foods were consumed frequently. These patterns mirror national dietary surveys, which show increasing energy intake from refined carbohydrates and processed foods among Thai children [2, 5, 18]. Similar trends have been reported internationally, where school-aged children’s diets are shaped by the surrounding food environment and family practices. Evidence indicates that improving the school food environment, through healthier food availability, canteen standards, or restrictions on sugary snacks, can positively influence children’s dietary choices and nutrient intake [23, 24]. Furthermore, involving parents and caregivers in school-based interventions enhances program effectiveness and supports sustained dietary behavior change at home [25, 26]. Educational interventions alone are therefore unlikely to shift behaviors without simultaneous improvements to the school food environment and family engagement in promoting healthier practices.

A key strength of this study was the active involvement of teachers, who played a pivotal role in integrating animated videos and interactive activities into their lessons. This approach enhanced student engagement and demonstrated the feasibility of embedding nutrition education within existing curricula. The use of digital tools, including accessible online platforms, showed that blended learning approaches can complement traditional classroom teaching without requiring extensive additional resources. The positive reception from teachers and students underscores schools’ readiness to engage in health-promoting activities when supported by appropriate training and materials. Similar studies have shown that teacher-led digital interventions improve engagement and support the adoption of innovative health education methods in primary schools [8, 27].

Another strength of this study was its multi-stage sampling design, which combined purposive district selection with disproportionate stratified random sampling of schools. Random selection within districts helped minimize selection bias and ensured representation across different school sizes, enhancing comparability between intervention and control groups. Nevertheless, limitations should be acknowledged. The district-level assignment of the intervention introduced potential contextual influences on the outcomes, limiting the generalizability of the findings beyond the semi-urban context of Chiang Mai. The sample size was calculated at the individual level and did not fully account for clustering within schools, which could have slightly reduced statistical power. One intervention school withdrew before implementation, leading to the loss of 30 students. This attrition reduced the total sample size and contributed to slight demographic imbalances, including the minor age difference between groups at baseline. Although ethnic differences were also present, both groups lived in similar semi-urban environments, and their household food preparers shared comparable characteristics, suggesting these differences were unlikely to have significantly influenced nutrition-related behaviors [28, 29].

In addition, the short duration of the intervention limited the ability to assess whether it had a lasting impact on students’ dietary intake. Dietary intake data were collected descriptively to provide contextual information about eating behaviors, but no statistical comparisons were performed between groups. As dietary habits often require consistent, long-term reinforcement to change, extended follow-up would be needed to determine whether future interventions could produce sustained improvements. Evidence from other studies supports the importance of intervention duration and comprehensiveness. For instance, a 12-week school-based program in Greece demonstrated significant and lasting reductions in BMI, energy intake, and fat intake, along with increased consumption of healthier foods, both in the short term (15 days) and long-term (12 months) follow-up [30]. A systematic review of European school interventions found that multi-component, longer-term strategies are more effective in improving dietary behaviors and BMI than short-term or education-only programs [31]. Similarly, a five-month program in Korean primary schools improved nutrition knowledge, dietary behaviors, and reduced body fat, highlighting the importance of sustained and reinforced educational efforts for promoting healthier eating among children [32].

Future research should prioritize strategies to enhance school participation and retention to minimize attrition and demographic imbalances. Although early engagement with schools and teachers was undertaken in this study, one school ultimately withdrew before implementation. This suggests that additional strategies, such as providing ongoing communication, administrative support, and addressing resource constraints, may be needed to maintain participation. Ensuring balanced baseline characteristics through stratified randomization would further improve comparability between intervention and control groups. Longer-term interventions with follow-up assessments are also essential to capture sustained impacts on dietary behaviors. Incorporating multi-component strategies, including parental involvement, experiential learning, and improvements to the school food environment, can strengthen intervention effectiveness and promote lasting behavior change.

## Conclusion

This study found no significant effects of a nine-week, school-based nutrition education intervention on students’ nutrition knowledge, attitudes, or behaviors. Nonetheless, the implementation process demonstrated that teacher-led, classroom-based nutrition education is feasible and well accepted in primary schools. The experience gained from this study provides a basis for refining future programs and evaluation methods. Longer-term and multi-component interventions that include parental and community involvement are recommended to achieve sustained improvements in children’s dietary behaviors and nutritional outcomes.

## Data Availability

The data underlying this study contain potentially identifiable information about student participants and cannot be shared publicly due to ethical restrictions imposed by the research ethics committee. De-identified data may be made available from the corresponding author upon reasonable request for researchers who meet the criteria for access to confidential data.

## Acknowledgements

The authors sincerely thank all participating students and teachers for their cooperation. We also acknowledge the Thailand Creative and Design Center and the Primary Educational Service Area Office 4 in Chiang Mai for their valuable support during fieldwork. The authors are grateful to Guido Lüchters for his helpful recommendations regarding statistical analyses. All authors reviewed and approved the final version of the manuscript.

## Supporting information

**S1 Table. Baseline information of household primary food preparers.** ^a^BMI: body mass index (underweight <18.5 kg/m²; normal weight 18.5–22.9; overweight 23.0–24.9; obesity class I 25.0–29.9; obesity class II ≥30.0); ^b^THB: Thai Baht, 1 THB ≈ 0.03 USD during the study period; ^c^Multiple places refers to respondents who reported purchasing meals from more than one source (e.g., food shops/restaurants, supermarkets/convenience stores, and/or markets).

**S2 Table. Energy and nutrient intakes per day based on 24HDR (presented with median (P25, P75).** ^a^The daily recommendation is based on the Thai Dietary Reference Intakes 2020, Department of Nutrition, Ministry of Public Health, Thailand; ^b^Percent distribution of carbohydrate: protein: fat = 45-65: 10-15: 30; ^c^≤ 10% of the total energy per day; ^d^≤ 300 mg per day; ^e^Daily dietary fiber requirement = age in years + 5 (for this study, 9–12 years = 14–17 g/day); ^f^≤ 5% of the total energy per day; 24HDR: 24-hour dietary recall; g: grams; kcal: kilocalories; RAE: retinol activity equivalent.

**S3 Table. Food consumption in grams per day based on semi-FFQ (eaters only).** FFQ: semi-food frequency questionnaire; %: percentage of children who consumed the food item (eaters only).

**S4 Table. Effectiveness of the nutrition education intervention on KAB scores adjusted for student covariates and parent factors.** All models were adjusted for students’ age, sex, ethnicity, and daily allowance, with a random intercept for school. Parental covariates (relation to child, education, occupation, income, age, sex, BMI, and meal preparation behavior) were included simultaneously in the model because variance inflation factors (all VIF < 2) indicated no problematic multicollinearity. The group × time coefficient represents the intervention effect.

**S5 Table. Effectiveness of nutrition education intervention on KAB scores adjusted for student covariates and 24HDR nutrient blocks.** All models adjusted for student age, sex, ethnicity, and daily allowance, with a random intercept for school. Dietary intake variables were analyzed in five separate blocks: Block 1, energy (kcal); Block 2, macronutrients (carbohydrate (g), protein (g), fat (g), saturated fatty acids (g)); Block 3, minerals (cholesterol (mg), calcium (mg), iron (mg), zinc (mg)); Block 4, vitamins and fiber (vitamin A (RAE), thiamin (mg), riboflavin (mg), vitamin C (mg), dietary fiber (g)); and Block 5, sodium (mg) and sugar (g).

**S6 Table. Effectiveness of the nutrition education intervention on KAB scores adjusted for student covariates and FFQ food groups.** All models were adjusted for students’ age, sex, ethnicity, and daily allowance, with a random intercept for school. FFQ dietary intake variables (rice and related products, fruits, vegetables, eggs and meats, milk, sugary drinks, and snacks) were included simultaneously in one model because multicollinearity diagnostics (all VIF < 2) indicated no problematic correlations. In contrast to Supplementary Table S5, where nutrient intakes from the 24HDR were highly correlated and therefore analyzed in separate blocks, the FFQ food group variables were analyzed together to provide mutually adjusted estimates. The group × time coefficient represents the intervention effect.

